# Metabolomic Alterations in Crimean Congo Hemorrhagic Fever Explored Through a Nationwide Study Using NMR Spectroscopy

**DOI:** 10.1101/2023.08.16.23294178

**Authors:** Oktay Göcenler, Kerem Kahraman, Derya Yapar, Yaren Kahraman, Cengizhan Büyükdağ, Gülen Esken, Serena Ozabrahamyan, Tayfun Barlas, Yüksel Karadağ, Aysel Kocagül Çelikbaş, Füsun Can, Nurcan Baykam, Mert Kuşkucu, Önder Ergönül, Çağdaş Dağ

**Affiliations:** Nanofabrication and Nanocharacterization Center for Scientific and Technological Advanced Research (n2STAR), Koç University, İstanbul, Türkiye; Hitit University, School of Medicine, Çorum, Türkiye; Department of Infectious Diseases and Clinical Microbiology, Çorum Erol Olçok Training and Research Hospital, Çorum, Türkiye; Koç University Isbank Center for Infectious Diseases (KUISCID), Koc University, Istanbul, Türkiye; The Ohio State University Wexner Medical Center, Columbus, USA; Koç University School of Medicine, Department of Medical Microbiology, Istanbul, Türkiye; Koç University School of Medicine, Department of Clinical Microbiology and Infectious Diseases, Istanbul, Türkiye

## Abstract

Crimean-Congo Hemorrhagic Fever (CCHF) is a severe tick-borne viral disease with high mortality rates and significant public health implications. Despite its global prevalence, the mechanisms underlying its pathogenesis remain poorly understood, and effective diagnostic and therapeutic tools are limited. Metabolomics, as a powerful tool for exploring host-pathogen interactions, offers a promising avenue for identifying biomarkers and elucidating disease mechanisms. In this study, we investigated the metabolic alterations in CCHF patients using non-targeted metabolomics to enhance understanding of disease pathogenesis, improve diagnostic capabilities, and identify potential therapeutic targets. A nationwide analysis was conducted on the blood serum of 29 CCHF patients and 10 healthy controls, employing Nuclear Magnetic Resonance (NMR) spectroscopy. Serum samples were collected over four consecutive days, and metabolic profiling was performed using Partial Least Squares Discriminant Analysis (PLS-DA) and Variable Importance in Projection (VIP) scoring to identify key metabolic pathways and compounds. Significant disruptions in metabolic pathways were observed in CCHF patients, particularly in purine and pyrimidine metabolism, the TCA cycle, and redox-related processes. Elevated levels of metabolites such as S-adenosyl homocysteine (SAH), guanosine triphosphate (GTP), inosine monophosphate (IMP), adenosine monophosphate (AMP), carnosine, 2’-deoxyuridine, nicotinamide adenine dinucleotide phosphate (NADP+), and maleate were identified. These metabolites demonstrated potential as biomarkers for disease severity and progression, with distinct metabolic profiles observed between moderate and severe cases. This study provides the first comprehensive metabolomic analysis of CCHF, highlighting critical metabolic pathways disrupted during infection. The findings underscore the utility of NMR-based metabolomics for identifying biomarkers that facilitate early diagnosis, prognosis, and therapeutic development. These results pave the way for future research to validate the identified biomarkers and explore targeted treatment strategies to improve patient outcomes in this severe viral infection.

## 1. Introduction

Crimean-Congo Hemorrhagic Fever (CCHF) is a severe viral infection with substantial implications for public health in a wide range of countries in Asia, Africa, Southern Europe and the Middle East. CCHF is caused by CCHF virus (CCHFV) belonging to the genus of Orthonairovirus, and the family of Nairoviridae being one of deadliest viruses of its kind with reported mortality rate of 3-30% (1). CCHFV is transmitted to humans through tick bites of infected ticks, contact with blood or tissues of infected livestock, or contact with infected patients (2). The exact course of pathogenesis of CCHF is not clearly known however it is divided into four phases: incubation, pre-haemorrhagic, haemorrhagic and convalescence [2]. CCHF is classified as a severe hemorrhagic fever with a short incubation period of 1-3 days although longer incubation periods have been documented [3]. The onset of infection is often sudden and includes symptoms, fever, diarrhea, vomiting, nausea, myalgia, back and abdominal pain followed by an hemorrhagic phase where severe bruises, uncontrollable bleeding at the body orifices are observed and in severe cases, deterioration of kidneys, liver and lungs [4]. Deaths associated with the infection mostly occur between 5-14 days from the start of the viremic phase [5]. In terms of treatment, early hospitalizations and early administration of therapeutics are shown to reduce both severity and mortality of CCHF [6]. Hence, lack of early detection of CCHF is one of the leading factors causing the particular high mortality rate of CCHF.

There are several challenges of diagnosing CCHF infection particularly before the hemorrhagic phase of the infection and patients who are not suspected of being bitten by infected ticks or contacted with infected livestock. CCHF is comparatively uncommon in specific regions, which may lead healthcare providers to initially overlook it as a potential diagnosis. Likewise, there are documented cases of difficulties diagnosing CCHF, both because of the latter but also the absence of a universally applicable diagnostic kit for surveillance and diagnosis of all CCHFV strains [7]. Standard blood tests such as hemogram, biochemical analysis and physical examination at the beginning of hospitalization is applied on all patients who are suspected to be infected with CCHF although the results are often relevant for short term prognostic factors as biochemical values change often quickly. For more comprehensive and standard diagnostic methods, viral antigen and nucleic acid amplification tests are employed [8]. The initial symptoms of CCHF, such as fever, headache, and muscle aches, are rather non-specific hence diagnosing CCHF early by differential diagnosis can be difficult [5]. However biomarkers, essentially biomolecules, may provide a measure of specific diseases or their stages due to their varying concentrations. Bio markers are instrumental in diagnosing and monitoring the progression of viral infections. The associated changes in their levels, often indicative of the disease, are typically attributed to the host’s immune reaction and the disturbance of key biochemical routes in reaction to the infectious process. In this sense, omics studies and biomarkers could be used for both analysis and diagnosis of CCHF and for characterizing better treatment strategies of hospitalized patients swiftly is crucial to pinpoint optimal treatment strategies for treating CCHF [9].

So far there is only one omics study which investigates host-viral response and pathogenesis of CCHF utilizing transcriptomics and proteomics methods [10]. Led by this gap in the literature, we conducted a nationwide analysis of metabolomes of patients hospitalized due to CCHF. Turkey has over 10,000 cases of CCHF with an average fatality rate of 5%, making it a critical public health concern affecting people living in rural areas as ticks are widespread in these regions [11]. In our present investigation, we employed Nuclear Magnetic Resonance (NMR) spectroscopy exploratory metabolomics to investigate the overall temporal variations in plasma metabolites during a seasonal outbreak of CCHF infection. We employed PLS-DA statistical analysis of the blood serum metabolome of CCHF patients and categorized certain metabolomes linked to metabolic dysregulation caused by CCHF. Preliminary results from our study suggest that specific metabolic markers can be identified in the serum of CCHF patients pointing to metabolic dysregulation, which may allow for earlier diagnosis and more targeted treatment strategies. Additionally, being the first study to categorize alterations of patient metabolome during CCHF viremic phase may be valuable for efforts to develop therapeutics or targeted treatment strategies to reduce the severity and high mortality rate of CCHF.

## 2. Materials and methods

### Monitoring of CCHF diagnosed patients and collection of the samples from the patients

Patients who were diagnosed with CCHF were selected from Çorum Hitit University Hospital. The first sample collection was conducted in April 2022, and blood samples were collected daily using Ethylenediaminetetraacetic acid (EDTA) tubes. In current metabolomics studies, opinions regarding the appropriate determination of sample size can considerably vary. However, numerous statistical analyses have underscored that a substantial sample size, for achieving meaningful results, around 30 samples [12]. In this study, serum specimens were procured from 29 patients (n=29) diagnosed with CCHFV infection, as well as from 10 healthy control group. CCHF patients also admitted to the hospital with an infection diagnosis were subclassified into two categories based on their blood test values and symptoms: moderate (n=24) and severe (n=5). Four blood serum samples from patients each consecutive day (n=116 samples) and a single sample from the control group (n=10 samples) was taken. These blood samples were then subjected to centrifugation for 5 minutes at 3000g to separate the sample into plasma, white blood cell, and red blood cell phases. The plasma phase was extracted and subjected to metabolite extraction using multiple approaches: single methanol extraction, triple alcohol extraction, Methanol-chloroform, Acetone, Acetonitrile, and Ultrafiltration. Cold methanol-chloroform was chosen as the most effective extraction and used for extraction of all samples. Only the polar metabolites in the plasma were investigated, while proteins and apolar compounds were removed from the plasma samples. Overall in this study, we utilized a rigorous approach to sample collection, preparation, and analysis to investigate polar metabolites in the plasma of CCHF patients [13].

### Ethical Statement

The study was approved by Koç University Committee on Human Research (01.12.2021/ 2021.436.IRB2.079) and all the procedures performed in this study involving human participants were in accordance with the ethical standards of the institutional research committee ethical standards. A written informed consent was obtained from all patients.

### NMR sample preparation

4 mL ice-cold methanol-chloroform (1:1) mixture was added to 2 mL serum for methanol-chloroform extraction. The mixture was vortexed for 30 seconds and incubated for 10 minutes on ice. After incubation the mixture was centrifuged at 4500g at 4°C for 30 minutes. The methanol phase was collected and dried using a vacuum concentrator. The dried samples were dissolved in 550 uL D_2_O based NMR sample solution (50 mM PBS (pH 7.4), 20mM NaCl, 1 mM DSS) for standardized sample preparation.

### NMR data collection, processing and statistical analysis

500 MHz Bruker Ascend magnet with BBO paired resonance probe and Avance NEO console was used for NMR data collection. 1D NOESY-presat (noesygppr1d) pulse sequence was used for data collection. Each NMR data spectrum is composed of 4K screening and 32K complex data points. Spectrum widths were set to 9615.4 Hz. Bruker Topspin 4.2.0 software was used for NMR data processing. Data was divided into 0.02 ppm data packages along with their normalization coefficients. The dataset, which comprises data packets with a resolution of 0.02 ppm, was analyzed using the MetaboAnalyst 5.0 online metabolomics statistical analysis software. Henceforth the data will be referred to as (Bin.x.xx [ppm]) data packets and the day of when sample was taken. All data points were normalized using the average centering normalization method. Following this normalization, the dataset underwent statistical analysis using Partial Least Squares Discriminant Analysis (PLS-DA). PLS-DA is a classification and discrimination technique based on the Partial Least Squares (PLS) regression method. This method is widely utilized to determine the differences between classes, particularly in high-dimensional and multivariate datasets. PLS-DA is a commonly employed method in analyzing complex biological systems, such as metabolomics studies. The VIP Projection variable importance score plot is a graph that is utilized to assess the outcomes of PLS-DA and determine the most significant variables in the analysis. VIP scores quantify the importance of each variable (e.g., metabolites) in classification and aid in identifying the most critical features. VIP scores are computed based on the contribution of each variable to the components (latent variables) in the PLS-DA model. The values begin at 1, and higher VIP scores indicate that the variable is more important for classification. Variables with VIP scores greater than 1 are generally deemed significant, although this threshold may vary in practice. The VIP score plot displays the VIP scores of the variables on the vertical axis, while the variables themselves or their indices are shown on the horizontal axis. This graph facilitates the identification of important variables visually and helps focus on the variables that require prioritization in the analysis. In the VIP score graph, the peaks at the relevant ppm values that make up the data packages (Bin.x.xx) have been examined in more detail and the metabolites to which they belong have been identified. For this operation, NMR spectra have been reopened, and the metabolites to which the peak in the relevant ppm region belongs have been determined using the Chenomx software. It is thought that some peaks might belong to metabolites not found in the database, and the molecules these peaks belong to have not been identified. Further investigation and characterization may be required to fully understand these unidentified peaks and their role in the overall metabolic profile, ensuring that the final analysis provides an accurate reflection of the biological system under investigation.

### Metabolomic pathway visualization

All metabolic pathways visualized using Metastate software Version BETA (https://metastate.bio) (Metastate Bio Inc.). Metastate algorithm employs the Kyoto Encyclopedia of Genes and Genomes (KEGG) database as its foundational input source. Software systematically retrieves details pertaining to biological pathways, chemical compounds, and molecular reactions of interest. Software curates and assembles a dynamic graphical representation of the data.

## 3. Results

In this study, 29 patients diagnosed with Crimean-Congo Hemorrhagic Fever (CCHF) and 10 healthy individuals serving as control subjects were included. The control group had a mean age of 50.1 years (range: 40–64 years), while the CCHF patient group had a slightly higher mean age of 50.5 years, with a wider age range of 22–77 years. A total of 126 blood samples were collected for analysis. Serum samples were prepared using methanol extraction, followed by Nuclear Magnetic Resonance (NMR) spectroscopy. The metabolite profiles obtained were subjected to comprehensive statistical analysis. This analysis incorporated data from all collected samples over four consecutive days and categorized them into moderate, severe, and control groups for comparative evaluation. Detailed procedures are described in the Materials and Methods section.

Partial Least Squares Discriminant Analysis (PLS-DA) was employed to develop a model incorporating the primary components (latent variables) of the dataset. The model focused on the first five components, which were pivotal for data classification and accounted for most of the dataset’s variance. The corresponding score plots, presented as two-dimensional graphs, illustrate the pairwise comparisons of these components (Figure 1a).

**Figure 1.**
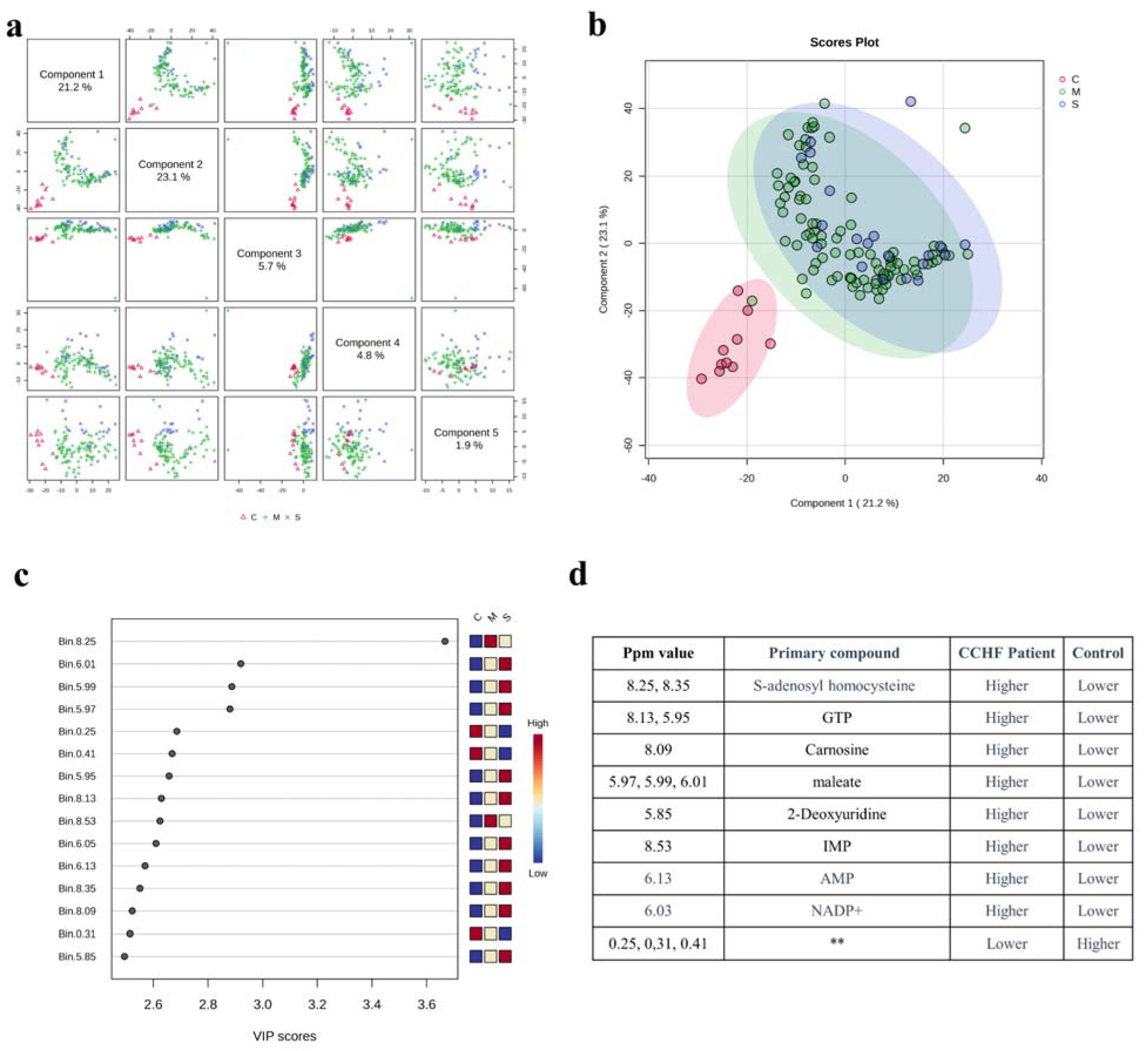
PLS-DA analysis of patient samples with severe and moderate infection level for four consecutive days with the control group (S: severe, M: moderate, C: Control) a. Matching score plots for the first five components of the PLS-DA analysis of all samples. b. partial Least Squares Discriminant Analysis score plot. c. Variable importance score plot in PLS-DA VIP Projection. d. corresponding compounds in detected increases on the spectra. Component 1: accuracy=0.817, (R2=0.246, Q2=0.142), component 2: accuracy=0.833, (R2=0.394, Q2=0.210), component 3: accuracy=0.809, (R2=0.545, Q2=0.225), component 4: accuracy=0.777, (R2=0.644, Q2=0.250), component 5: accuracy=0.769, (R2=0.782, Q2=0.205)

The PLS-DA score plot (Figure 1b) clearly differentiates between healthy and diseased individuals. PLS-DA, well-suited for handling high-dimensional data, effectively identifies the key variables responsible for distinguishing health states. This approach revealed a distinct separation based on variations and correlations within the biomarker data. To further explore the critical contributors to this separation, the Variable Importance in Projection (VIP) scores were analyzed. The PLS-DA VIP score plot (Figure 1c) highlights the variables with the most significant influence on the discrimination process, with the top 15 variables identified. VIP scores provide insights into the relevance of each variable, aiding in the identification of biomarkers that significantly differ between healthy and diseased states.

Figure 1c demonstrates that some data buckets exhibit lower concentration values (indicated in blue) in the control group, which progressively increase over time. Furthermore, the identified compounds displayed a marked increase in concentration, as corroborated by enhanced signals from their respective data buckets (Figure 1d).

Statistical analysis was performed across three distinct groups, incorporating samples collected over four days from both patient groups. While patient samples were successfully distinguished from the control group, no significant differences were observed between the moderate and severe disease groups. Key metabolites found to be elevated in the patient groups compared to controls included SAH, GTP, carnosine, maleate, 2-deoxyuridine, IMP, AMP, and NADP+.

As a secondary approach to data analysis, metabolite profiles were evaluated using samples collected on days 1 and 2 of hospitalization, representing the early stages of infection. Patients were categorized into severe and moderate groups based on clinical severity, while blood samples from healthy individuals were used as the control group for statistical comparisons (Figure 2). This focused analysis aimed to investigate metabolite changes during the initial phase of the disease. In addition to compounds identified in the initial analysis, novel metabolites with significant increases on days 1 and 2 were detected, suggesting their potential involvement in the pathogenesis of Crimean-Congo Hemorrhagic Fever (CCHF).

**Figure 2.**
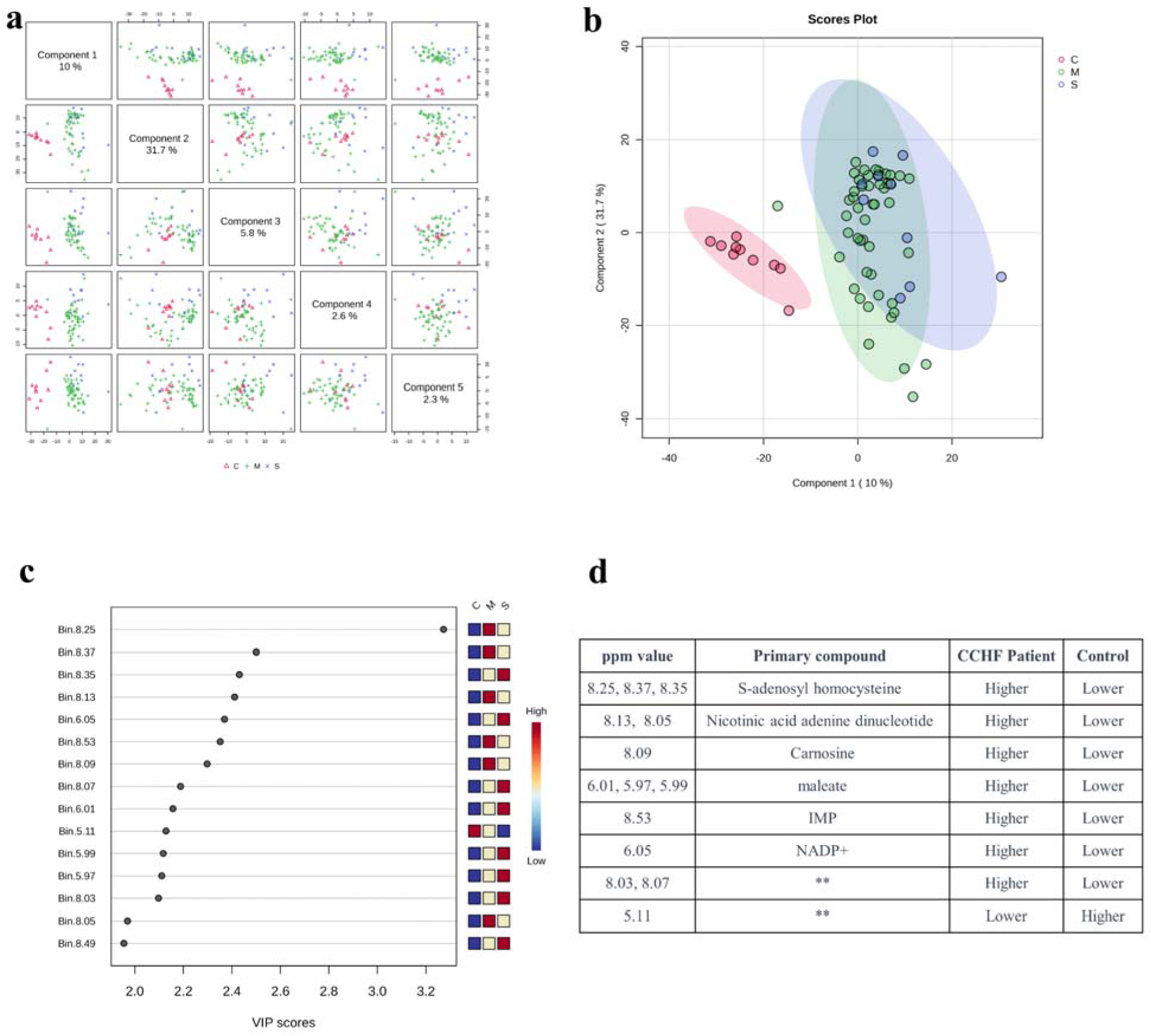
PLS-DA analysis of patient samples with severe and moderate infection level for first two days with the control group matching score plots for the first five components of the PLS-DA analysis of samples from day 1 and day 2 as labeled mild (M), severe (S) and compared with the control group (C). b, partial Least Squares Discriminant Analysis score plot. c, Variabl importance score plot in PLS-DA VIP Projection. d, corresponding compounds in detected increases on the spectra. Component 1: accuracy=0.0837, (R2=0.585, Q2=0.417), component 2: accuracy=0.821, (R2=0.627, Q2=0.433), component 3: accuracy=0.793, (R2=0.804, Q2=0.352)

A comprehensive metabolomic analysis of blood serum from CCHF patients revealed distinct metabolite patterns corresponding to infection severity. Partial Least Squares Discriminant Analysis (PLS-DA) score plots and Variable Importance in Projection (VIP) scores (Figure 3) highlighted notable differences in metabolite profiles between severe and moderate cases. In severe CCHF infections, metabolites such as AMP, IMP, and NAAD were significantly elevated in the serum, whereas these compounds were less prominent in samples from patients with moderate infection. Conversely, GTP was markedly increased in the moderate infection group but was not significantly detected in the severe group.

**Figure 3.**
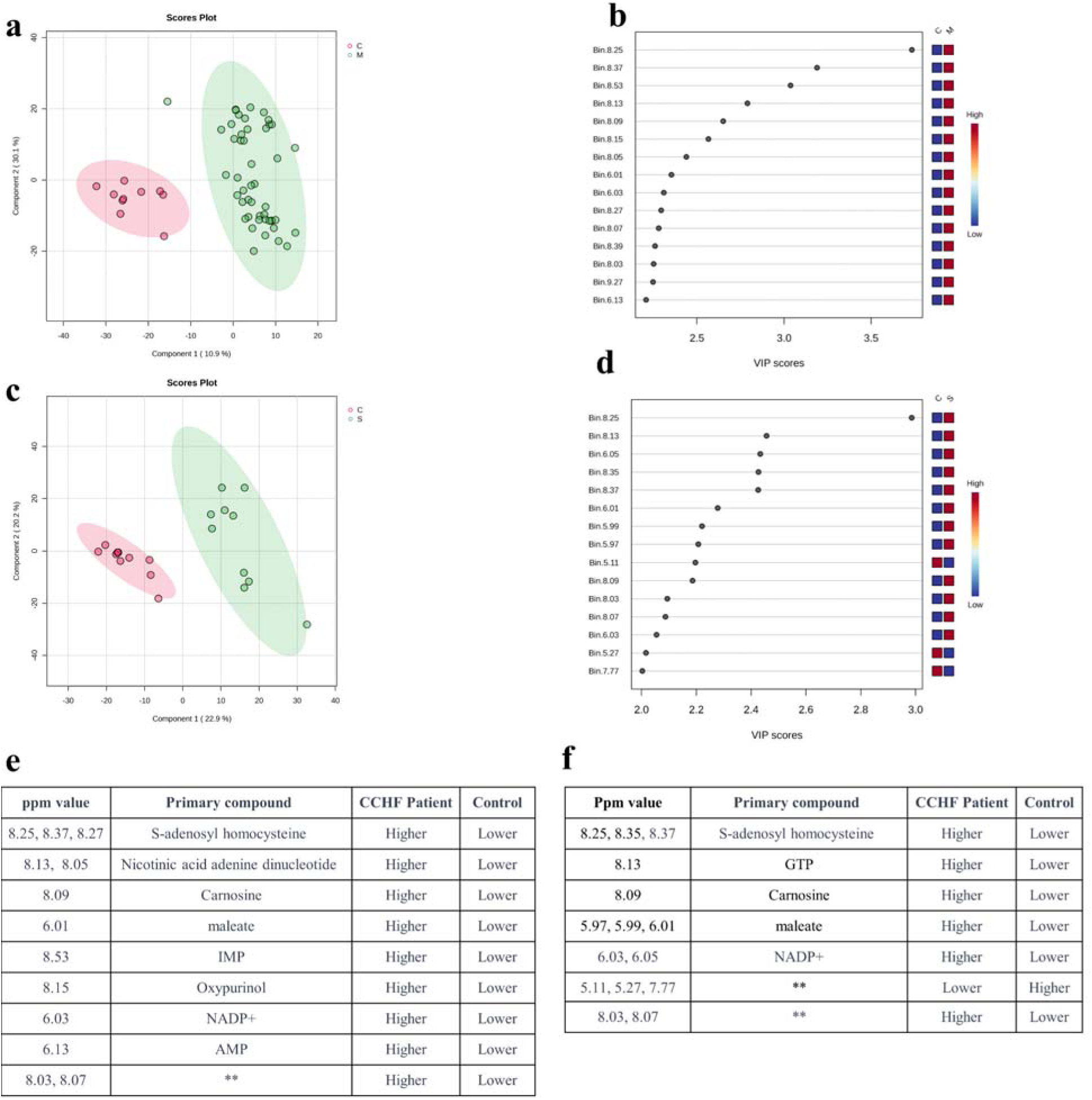
PLS-DA analysis of patient samples with moderate infection level for first two days with the control group and PLS-DA analysis of patient samples with severe infection level for first two days with the control group (S:severe, M: moderate, C:Control) a, partial Least Squares Discriminant Analysis score plot. Component 1: accuracy=0.983, (R2=0.821, Q2=0.739), component 2: accuracy=0.983, (R2=0.860, Q2=0.790), component 3: accuracy=0.983, (R2=0.954, Q2=0.829) b, Variable importance score plot in PLS-DA VIP Projection. c, partial Least Squares Discriminant Analysis score plot. Component 1: accuracy=0.950, (R2=0.857, Q2=0.702), component 2: accuracy=1, (R2=0,938, Q2=0.797), component 3: accuracy=1, (R2=0.982, Q2=0.824) d, Variable importance score plot in PLS-DA VIP Projection. e, corresponding compounds in detected increases on the spectra. f, corresponding compounds in detected increases on the spectra.

Overall, these findings highlight substantial metabolic differences between moderate and severe CCHF infections, pointing to critical pathways potentially involved in CCHF pathogenesis. These results underscore the importance of early-stage metabolite profiling to enhance our understanding of disease mechanisms and identify potential biomarkers for severity stratification.

## 4. Discussion

In this study, we present the first non-targeted metabolomics analysis of Crimean-Congo Hemorrhagic Fever (CCHF) to enhance the understanding of its pathogenesis, improve diagnostic capabilities, and aid in the development of potential therapeutic interventions. Using Nuclear Magnetic Resonance (NMR) spectroscopy, we identified significant increases in key metabolites, including S-adenosyl homocysteine (SAH), guanosine triphosphate (GTP), carnosine, maleate, 2’-deoxyuridine (2’-dU), inosine monophosphate (IMP), adenosine monophosphate (AMP), and nicotinamide adenine dinucleotide phosphate (NADP+), in the blood serum of CCHF patients. These findings suggest that these metabolites may play critical roles in the pathogenesis of CCHF and serve as important biomarkers for early detection and monitoring of disease progression.

Metabolomics has emerged as a powerful tool for studying host-pathogen interactions by revealing metabolic alterations induced by viral infections. To date, only one prior omics study utilizing mass spectrometry (MS) has investigated the interaction between the host and CCHF virus (CCHFV). Both NMR and MS are widely used in metabolomics, with each method offering distinct advantages depending on the research objectives. NMR spectroscopy is particularly suitable for non-targeted metabolomics due to its superior reproducibility, minimal sample preparation, and ability to analyze complex biofluids. It also provides consistent spectra across different instruments and laboratories, making it highly reliable for diagnostic and prognostic studies. On the other hand, MS offers higher sensitivity and is often preferred for targeted metabolomics studies.

The utility of NMR metabolomics has been demonstrated in the analysis of host metabolic changes induced by various viral infections, including HIV, dengue virus (DENV), and chikungunya virus. Moreover, metabolomics approaches have been successfully applied to study other viral hemorrhagic fevers, such as Ebola, Marburg, and dengue, yielding promising results for early diagnosis and prognosis prediction. Similarly, our findings underscore the potential of NMR metabolomics in elucidating the complex metabolic interactions between CCHFV and its host. The observed metabolic changes in CCHF patients offer new insights into the mechanisms of disease pathogenesis and highlight potential pathways for targeted therapeutic interventions.

By focusing on metabolite profiles in CCHF, this study demonstrates the value of NMR-based metabolomics in addressing the gaps in knowledge surrounding this severe disease. Our findings not only advance the understanding of CCHF pathogenesis but also provide a foundation for further research aimed at identifying effective biomarkers and therapeutic targets. These results emphasize the critical role of metabolomics in uncovering host-pathogen dynamics and improving clinical outcomes in viral infections.

Among the notable metabolites detected, S-adenosyl homocysteine (SAH) emerged as a significant intermediate in the metabolic pathways of CCHF patients. SAH, a precursor to homocysteine and adenosine, serves as the substrate for the enzyme SAH hydrolase, a critical component of the S-adenosylmethionine (SAM/AdoMet) regeneration cycle (Figure 4a). Elevated SAH levels were observed in CCHF patients early during hospitalization, underscoring its potential relevance in the disease’s pathogenesis. This observation aligns with findings from a genomic study linking Methylenetetrahydrofolate reductase (MTHFR) polymorphisms to a predisposition for milder forms of CCHF [18]. Since MTHFR plays a central role in folate metabolism and methylation processes, these results suggest that disruptions in methylation pathways during the viremic phase could serve as prognostic indicators for CCHF severity.

**Figure 4.**
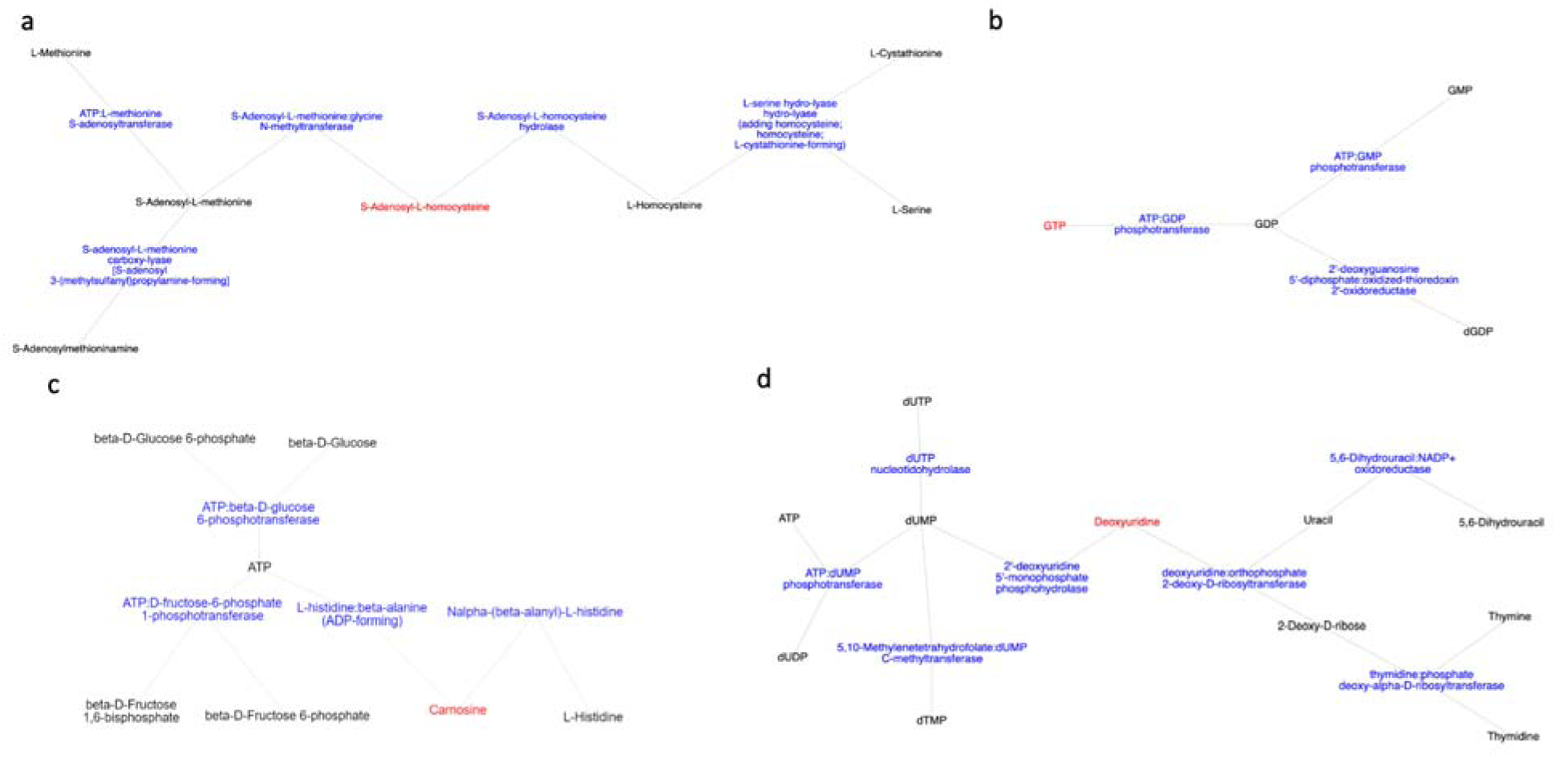
Main metabolic pathways of metabolites detected by metabolomics approach a. S-Adenosyl-L-homocysteine and nearest neighbor in Cysteine and methionine metabolism b. GTP and nearest neighbor in Purine metabolism c. Carnosine and nearest neighbor in Histidine metabolism d. Deoxyuridine and nearest neighbor in Pyrimidine metabolism

SAM is a key methyl donor in various cellular methylation reactions, including those involved in 5’ RNA capping, a process critical for viral replication and transcription [19]. Evidence from other viral families, such as flaviviruses [20], and Ebola, whose L protein exhibits methyltransferase activity, indicates that methylation mechanisms can influence RNA cap structure and internal adenosine-2’-O-methylation [21]. These parallels highlight the potential importance of methylation pathways in CCHF pathogenesis. The additional complexity of RNA methylation in some viral families suggests that SAM-related domains within viral replication complexes could represent promising therapeutic targets.

Our findings emphasize the need for further research into SAM-related pathways to unravel their implications for viral replication and host-pathogen interactions in CCHF. These pathways not only provide insight into disease mechanisms but also present opportunities for the development of targeted therapeutic interventions, particularly those aimed at disrupting key methylation processes critical for viral survival.

GTP, AMP, and inosine monophosphate (IMP) are key nucleotides involved in purine metabolism, essential for energy production and nucleic acid synthesis (Figure 4b). Elevated levels of GTP and IMP are well-documented indicators of viral infections, particularly among viruses in the *Orthornavirae* kingdom. In the context of CCHF, the increase in these purine metabolites aligns with the known mechanism of action of broad-spectrum antivirals like Ribavirin. As a nucleotide analog, Ribavirin targets the substrate-binding site of the IMPDH enzyme, thereby reducing RNA synthesis in infected cells by downregulating GTP synthesis [22]. Monitoring these metabolites could provide valuable insights into the efficacy of antiviral treatments. Additionally, quantifying shifts in GTP and IMP concentrations offers a potential strategy to optimize therapeutic interventions in clinical trials, enhancing the effectiveness of antiviral agents targeting purine metabolism [23].

Carnosine (β-alanyl-L-histidine) (Figure 4c), a dipeptide with antioxidant, anti-glycation, and anti-inflammatory properties, was also significantly elevated in CCHF patients. Predominantly found in skeletal muscle and the brain, increased carnosine levels may reflect a compensatory response to oxidative stress, infection-induced cell death, or muscle mass reduction due to hospitalization. Carnosine has demonstrated antiviral potential against Zika, dengue, and SARS-CoV-2, reducing viral replication and alleviating symptoms. While its therapeutic utility against CCHF remains to be established, these findings highlight the need for further investigation into its potential as a treatment option.

2’-Deoxyuridine (2’-dU), an intermediate in thymidylate synthesis (Figure 4d), also showed significant elevation in CCHF patients. This metabolite, a precursor for DNA synthesis, plays a role in antiviral therapies, such as Edoxudine, which target DNA viruses [24–26]. Variants of 2’-dU, such as BVDU, have shown efficacy against Herpes simplex virus type 1 (HSV-1) and varicella-zoster virus (VZV) [27]. While 2’-dU is typically associated with DNA virus activity, its elevated levels in CCHF patients may suggest broader disruptions in pyrimidine metabolism, potentially linked to the metabolic dysregulation observed in this study.

Nicotinamide adenine dinucleotide phosphate (NADP+) serves as a critical cofactor in enzymatic reactions, primarily within the pentose phosphate pathway, where it supports fatty acid synthesis and the regeneration of reduced glutathione. Its role in redox reactions also underscores its importance in the antioxidant defense system. Elevated NADP+ levels in CCHF patients could reflect increased metabolic activity related to energy production and nucleotide synthesis. This hypothesis is further supported by the observed overlap in the elevations of NADP+, GTP, IMP, and AMP, indicating a coordinated upregulation of energy metabolism pathways. The significant increase in these metabolites in severe cases (Figure 3) underscores their potential relevance in disease progression and severity.

Maleate (cis-butenedioic acid), a dicarboxylic acid and trans-isomer of fumaric acid, plays a role in nicotinate and nicotinamide metabolism [29]. Elevated maleate levels in CCHF patients suggest underlying metabolic disruptions. Previous studies have demonstrated that maleate can inhibit the tricarboxylic acid (TCA) cycle, lower ATP levels, and impair enzymatic activity, leading to systemic metabolic imbalances. These findings warrant further investigation into the potential implications of maleate in the pathophysiology of CCHF [30].

In summary, these metabolomic findings reveal significant disruptions in purine, pyrimidine, and energy metabolism pathways in CCHF patients. Metabolites such as GTP, IMP, AMP, carnosine, 2’-dU, NADP+, and maleate provide valuable insights into the pathogenesis of the disease and highlight potential biomarkers for disease progression and therapeutic targets.

## Conclusion

In this study, we conducted a nationwide analysis of blood serum metabolites from patients hospitalized due to Crimean-Congo Hemorrhagic Fever (CCHF), employing Nuclear Magnetic Resonance (NMR) spectroscopy as a novel approach to investigate host-pathogen interactions and the pathogenesis of CCHFV. Our findings revealed significant alterations in several key metabolites, including S-adenosyl homocysteine (SAH), guanosine triphosphate (GTP), inosine monophosphate (IMP), adenosine monophosphate (AMP), 2’-deoxyuridine (2’-dU), nicotinamide adenine dinucleotide phosphate (NADP+), carnosine, and maleate. Notable changes were observed in pathways related to the TCA cycle, nucleic acid synthesis, and redox-associated coenzymes, highlighting the systemic metabolic impact of CCHFV infection.

The elevation of metabolites such as SAH underscores the potential impact of methylation processes on CCHF pathogenesis and prognosis, linking viral replication to host epigenetic regulation. Similarly, the observed disruptions in purine metabolism, particularly the increased levels of GTP and IMP, suggest potential therapeutic implications, as these pathways are targeted by antiviral agents like Ribavirin. The role of other metabolites, such as carnosine and maleate, indicates a systemic response to oxidative stress and cellular damage, providing further insight into the host’s metabolic adaptation during infection.

Our findings align with prior metabolomics studies on other viral hemorrhagic fevers and highlight the utility of NMR-based metabolomics for uncovering biomarkers that may facilitate early diagnosis, prognosis prediction, and therapeutic development. The distinct metabolic profiles observed between moderate and severe CCHF cases further emphasize the relevance of these biomarkers in stratifying disease severity and monitoring treatment efficacy.

This study paves the way for future investigations to validate these biomarkers in larger cohorts and explore the therapeutic potential of targeting the identified pathways. By advancing the understanding of CCHF pathogenesis, our results contribute to the development of more effective diagnostic and therapeutic strategies, ultimately improving patient outcomes in this severe viral infection.

## Author Contributions

**Ça**ğ**da**ş **Da**ğ: Conceptualization, Methodology, Supervision, Funding acquisition, Writing-Reviewing and Editing, Writing-Original draft preparation **Kerem Kahraman**: Investigation, Formal analysis, Visualization, Writing-Reviewing and Editing **Oktay Göcenler:** Investigation, Formal analysis, Writing-Original Draft, Visualization **Derya Yapar:** Investigation **Yaren Kahraman:** Investigation, Formal analysis **Cengizhan Büyükda**ğ: Formal analysis, Writing-Original Draft **Gülen Esken:** Methodology, Investigation **Serena Ozabrahamyan:** Methodology, Investigation **Tayfun Barlas:** Investigation **Yüksel Karada**ğ: Investigation **Aysel Kocagül Çelikba**ş: Investigation **Füsun Can:** Supervision, Investigation **Nurcan Baykam:** Supervision, Investigation **Mert Ku**ş**kucu:** Supervision, Writing-Reviewing and Editing **Önder Ergönül:** Supervision, Funding acquisition, Conceptualization, Methodology

## Data Availability

All data produced in the present study are available upon reasonable request to the authors

## ACKNOWLEDGMENT

CD acknowledges support from TU□BI□TAK (Project No: 221S353). The authors acknowledge the use of the services and facilities of n^2^STAR-Koç University Nanofabrication and Nanocharacterization Center for Scientific and Technological Advanced Research. The authors gratefully acknowledge use of the services and facilities of the Koc University Is Bank Infectious Disease Center (KUIS-CID). We are also immensely grateful to Fırat Kahya, Boran Saruhan and Oğuzcan Ünver for constructive feedback and for their comments.

## References

[1] Belhadi, D., Baied, M. E., Mulier, G., Malvy, D., Mentré, F., & Laouénan, C. (2022). The number of cases, mortality and treatments of viral hemorrhagic fevers: A systematic review. PLOS Neglected Tropical Diseases, 16(10), e0010889. 10.1371/journal.pntd.0010889

[2] Hoogstraal H. (1979). The epidemiology of tick-borne Crimean-Congo hemorrhagic fever in Asia, Europe, and Africa. Journal of medical entomology, 15(4), 307–417. 10.1093/jmedent/15.4.307

[3] Whitehouse, C. A. (2004). Crimean?Congo hemorrhagic fever. Antiviral Research, 64(3), 145–160. 10.1016/j.antiviral.2004.08.001

[4] Elaldi, N., Bodur, H., Ascioglu, S., Celikbas, A. K., Özkurt, Z., Vahaboglu, H., Leblebicioglu, H., Yilmaz, N., Engin, A., Şencan, M., Aydin, K., Dokmetas, I., Çevik, M. A., Dokuzoğuz, B., Tasyaran, M. A., Öztürk, R., Bakir, M., & Uzun, R. (2009). Efficacy of oral ribavirin treatment in Crimean-Congo haemorrhagic fever: A quasi-experimental study from Turkey. Journal of Infection, 58(3), 238–244. 10.1016/j.jinf.2009.01.014

[5] Swanepoel, R., Gill, D. E., Shepherd, A. J., Leman, P. A., Mynhardt, J. H., & Harvey, S. (1989). The clinical pathology of Crimean-Congo hemorrhagic fever. Reviews of infectious diseases, 11 *Suppl 4*, S794–S800. 10.1093/clinids/11.supplement_4.s794

[6] Ergonul O. (2008). Treatment of Crimean-Congo hemorrhagic fever. Antiviral research, 78(1), 125–131. 10.1016/j.antiviral.2007.11.002

[7] Almayahi, Z. K., Kindi, H. A., Jabri, I. H. S. H. A., Shaqsi, N. H. K. A., Hattali, N. A., Hattali, A. A., Quyoodhi, B. A., & Dhuhli, K. A. (2022). Challenges in Diagnosis of Crimean-Congo Hemorrhagic Fever. Infectious Diseases in Clinical Practice, 30(2). 10.1097/ipc.0000000000001108

[8] Raabe V. N. (2020). Diagnostic Testing for Crimean-Congo Hemorrhagic Fever. Journal of clinical microbiology, 58(4), e01580–19. 10.1128/JCM.01580-19

[9] Mayne, E. S., George, J. A., & Louw, S. (2023). Assessing Biomarkers in Viral Infection. Advances in experimental medicine and biology, 1412, 159–173. 10.1007/978-3-031-28012-2_8

[10] Neogi, U., Elaldi, N., Appelberg, S., Ambikan, A. T., Kennedy, E. V., Dowall, S. D., Bagci, B., Gupta, S., Murillo, J. R., Akusjärvi, S. S., Monteil, V., Marko-Varga, G., Benfeitas, R., Banerjea, A. C., Weber, F., Hewson, R., & Mirazimi, A. (2022). Multi-omics insights into host-viral response and pathogenesis in Crimean-Congo hemorrhagic fever viruses for novel therapeutic target. eLife, 11. 10.7554/elife.76071

[11] Ak, Ç., Ergönül, Ö. & Gönen, M. A prospective prediction tool for understanding Crimean– Congo haemorrhagic fever dynamics in Turkey. Clin. Microbiol. Infect. 26, 123–e1 (2020).

[12] Nyamundanda, G., Gormley, I.C., Fan, Y. et al. MetSizeR: selecting the optimal sample size for metabolomic studies using an analysis based approach. BMC Bioinformatics 14, 338 (2013). 10.1186/1471-2105-14-338

[13] Markley, J. L., Brüschweiler, R., Edison, A. S., Eghbalnia, H. R., Powers, R., Raftery, D., & Wishart, D. S. (2017). The future of NMR-based metabolomics. Current opinion in biotechnology, 43, 34–40. 10.1016/j.copbio.2016.08.001

14. Emwas, A. M. (2015). The Strengths and Weaknesses of NMR Spectroscopy and Mass Spectrometry with Particular Focus on Metabolomics Research. In Methods in molecular biology (pp. 161–193). Springer Science+Business Media. 10.1007/978-1-4939-2377-9_13

[15] Munshi, S. U., Rewari, B. B., Bhavesh, N. S., & Jameel, S. (2013). Nuclear magnetic resonance based profiling of biofluids reveals metabolic dysregulation in HIV-infected persons and those on anti-retroviral therapy. PloS one, 8(5), e64298. 10.1371/journal.pone.0064298

[16] Shrinet, J., Shastri, J. S., Gaind, R., Bhavesh, N. S., & Sunil, S. (2016). Serum metabolomics analysis of patients with chikungunya and dengue mono/co-infections reveals distinct metabolite signatures in the three disease conditions. Scientific reports, 6, 36833. 10.1038/srep36833

[17] Wang, Z., Liang, H., Cao, H., Zhang, B., Li, J., Wang, W., Qin, S., Wang, Y., Xuan, L., Lai, L., & Shui, W. (2019). Efficient ligand discovery from natural herbs by integrating virtual screening, affinity mass spectrometry and targeted metabolomics. Analyst, 144(9), 2881–2890. 10.1039/c8an02482k

[18] Karakus, N., Duygu, F., Rustemoglu, A., & Yigit, S. (2022). Methylene-tetrahydrofolate reductase gene C677T and A1298C polymorphisms as a risk factor for Crimean-Congo hemorrhagic fever. *Nucleosides*, Nucleotides & Nucleic Acids, 41(9), 878–890. 10.1080/15257770.2022.2085296

[19] Byszewska, M., Śmietański, M., Purta, E., & Bujnicki, J. M. (2014). RNA methyltransferases involved in 5′ cap biosynthesis. RNA Biology, 11(12), 1597–1607. 10.1080/15476286.2015.1004955

[20] Brecher, M., Chen, H. S., Liu, B., Banavali, N. K., Jones, S., Zhang, J., Li, Z., Kramer, L. D., & Li, H. (2015b). Novel Broad Spectrum Inhibitors Targeting the Flavivirus Methyltransferase. PLOS ONE, 10(6), e0130062. 10.1371/journal.pone.0130062

[21] Valle, C., Martin, B., Ferron, F., Roig-Zamboni, V., Desmyter, A., Debart, F., Canard, B., Coutard, B., & Decroly, E. (2021). First insights into the structural features of Ebola virus methyltransferase activities. Nucleic Acids Research, 49(3), 1737–1748. 10.1093/nar/gkaa1276

[22] Tchesnokov, E. P., Bailey-Elkin, B. A., Mark, B. L., & Götte, M. (2020). Independent inhibition of the polymerase and deubiquitinase activities of the Crimean-Congo Hemorrhagic Fever Virus full-length L-protein. PLOS Neglected Tropical Diseases, 14(6), e0008283. 10.1371/journal.pntd.0008283

[23] Robins, R. K., Revankar, G. R., McKernan, P. A., Murray, B. K., Kirsi, J. J., & North, J. A. (1985). The importance of IMP dehydrogenase inhibition in the broad spectrum antiviral activity of ribavirin and selenazofurin. Advances in Enzyme Regulation, 24, 29–43. 10.1016/0065-2571(85)90068-8

[24] Rothan, H. A., Abdulrahman, A. Y., Khazali, A. S., Rashid, N. N., Chong, T. T., & Yusof, R. (2019). Carnosine exhibits significant antiviral activity against Dengue and Zika virus. Journal of Peptide Science, 25(8). 10.1002/psc.3196

[25] Saadah, L. M., Deiab, G. I. A., Al-Balas, Q., & Basheti, I. A. (2020). Carnosine to Combat Novel Coronavirus (nCoV): Molecular Docking and Modeling to Cocrystallized Host Angiotensin-Converting Enzyme 2 (ACE2) and Viral Spike Protein. Molecules, 25(23), 5605. 10.3390/molecules25235605

[26] Chon, J., Stover, P. J., & Field, M. S. (2017). Targeting nuclear thymidylate biosynthesis. Molecular Aspects of Medicine, 53, 48–56. 10.1016/j.mam.2016.11.005

27. De Clercq E. (2005). Potential clinical applications of the CXCR4 antagonist bicyclam AMD3100. Mini reviews in medicinal chemistry, 5(9), 805–824. 10.2174/1389557054867075

[28] Yen, Y. C., Kong, L. X., Lee, L., Zhang, Y. Q., Li, F., Cai, B. J., & Gao, S. Y. (1985). Characteristics of Crimean-Congo hemorrhagic fever virus (Xinjiang strain) in China. The American journal of tropical medicine and hygiene, 34(6), 1179–1182

[29] National Center for Biotechnology Information (2023). PubChem Compound Summary for CID 444266, Maleic Acid. Retrieved August 15, 2023 from https://pubchem.ncbi.nlm.nih.gov/compound/Maleic-Acid.

[30] Bergeron, M. G., Mayers, P., & Brown, D. T. (1996). Specific effect of maleate on an apical membrane glycoprotein (gp330) in proximal tubule of rat kidneys. American Journal of Physiology-renal Physiology, 271(4), F908–F916. 10.1152/ajprenal.1996.271.4.f908

